# How are sociodemographic factors associated with 2020/2021 seasonal influenza vaccination behavior under the COVID-19 pandemic?

**DOI:** 10.1101/2021.04.30.21256364

**Authors:** Takahiro Mori, Tomohisa Nagata, Kazunori Ikegami, Ayako Hino, Seiichiro Tateishi, Mayumi Tsuji, Shinya Matsuda, Yoshihisa Fujino, Koji Mori, the CORoNaWork project

**Author notes:** Correspondence author: Koji Mori, MD, PhD, Department of Occupational Health Practice and Management, Institute of Industrial Ecological Sciences, University of Occupational and Environmental Health, Japan, 1-1 Iseigaoka, Yahatanishi-ku, Kitakyushu 807-8555, Japan.

## Abstract

The 2020/2021 seasonal influenza vaccination was carried out under unique situations during the coronavirus disease 2019 (COVID-19) pandemic. Examining the factors affecting vaccine inoculation in a pandemic situation may provide valuable insights. The purpose of the current study was to investigate how the COVID-19 pandemic affected the 2020/2021 seasonal influenza vaccine intake. A cross-sectional study was conducted on workers aged 20–65 years on December 22–25, 2020, using data from an Internet survey. We set the presence or absence of 2020/2021 seasonal influenza vaccination as the dependent variable, and each aspect of sociodemographic factors, including gender, age, marital status, education, annual household income, and underlying disease, as independent variables. We performed a multilevel logistic regression analysis nested by residence. In total, 26,637 respondents (13,600 men, 13,037 women) participated, and a total of 11,404 individuals (42.8%) received the 2020/2021 influenza vaccine. Significantly more women than men were vaccinated, and the vaccination rate was higher among younger adults, married people, highly educated people, high-income earners, and those with underlying disease. The current results suggested that the relationship between seasonal influenza vaccination behavior and sociodemographic factors differed from the results reported in previous studies in terms of age. These findings suggest that, during the COVID-19 pandemic, young people may have become more aware of the risk of contracting influenza and of the effectiveness of the influenza vaccine. In addition, information interventions may have had a positive effect.

## Introduction

The Coronavirus disease 2019 (COVID-19) pandemic caused by Severe Acute Respiratory Syndrome Coronavirus 2, and the 2020/2021 seasonal influenza vaccination, which was started in the fall of 2020 when the development of the pandemic vaccine was urgently under way, share major symptomatic characteristics. Because the main complaints of influenza are fever and malaise, it is difficult to distinguish it from COVID-19 based on symptoms alone, and to diagnose COVID-19, nucleic acid amplification tests such as real-time reverse transcription–polymerase chain reaction (RT-PCR) or antigen tests are required.^1^ In Japan, although the number of infected people was small in comparison with other countries, the implementation system of tests for diagnosis was insufficient around the fall of 2020. If there was a strong suspicion of a new coronavirus infection after receiving instructions from a public health center or seeing an attending physician, the patient was mandated to undergo tests such as RT-PCR, the results of which were not immediately known.^2^ In addition, owing to the small number of infected people, those infected and their close contacts tended to be given special attention. In some cases, the workplace ordered them to remain at home longer than necessary. So, the 2020/2021 seasonal influenza vaccination was recommended for the purpose of preventing influenza, which is difficult to distinguish from COVID-19.^3^

Influenza vaccination is the most effective way to protect against seasonal influenza infections. Influenza is a common acute respiratory infection that can lead to hospitalization, and, in severe cases, death. Individuals at high risk of serious complications of influenza include people with asthma, chronic lung disease, diabetes mellitus, chronic heart disease, chronic kidney disease, stroke, obesity, central nervous system degeneration, immunosuppression, adults 65 years and older, children younger than 2 years old, and pregnant women.^4^ Many previous studies have reported that influenza vaccines have a range of benefits, including reducing the risk of influenza infection, hospitalization, and death, and inoculation is recommended, particularly for high-risk individuals.^4-7^ However, the influenza vaccination rate for individuals at high risk is 50% or less in many countries.^8^ In Japan, the inoculation rate for people aged 65 and over is less than 50%, and the inoculation rate for people under the age of 65 years is even lower.^8-10^

Kini et al. reviewed papers investigating the effects of age, gender, and race on seasonal influenza vaccination. Age was found to be directly related to vaccine intention, with vaccination rates increasing with age. Women were found to be more likely to receive the influenza vaccine, as were white populations.^11^ Schmid et al. reviewed papers from the 2005–2016 period covering the factors that influence influenza vaccine inoculation, and it has been reported that living alone, not being married, and having no diseases under treatment are associated with non-vaccination.^12^

In the 2020/2021 season, the expectation regarding the efficacy of the influenza vaccine included not only the prevention of influenza itself, but also the possibility of being treated as COVID-19.^3^ The vaccine itself is a widely used seasonal influenza vaccine, and the associated risk of adverse reactions is relatively well known. Investigating the factors influencing seasonal influenza vaccination during the COVID-19 pandemic may provide useful insights for improving the vaccination rate. The purpose of the current study was to investigate how the unique characteristics of the COVID-19 pandemic affected the 2020/2021 seasonal influenza vaccine intake.

## Materials and methods

### Study design

This cross-sectional study was conducted using data from a baseline study of a prospective cohort study called the Collaborative Online Research on the Novel-coronavirus and Work (CORoNaWork). This survey was conducted as a self-administered questionnaire by Internet research company Cross Marketing (Tokyo, Japan) to investigate the health status of workers from December 22 to December 26, 2020, when the third wave of the COVID-19 pandemic was circulating in Japan [13]. Details of the protocol have already been reported. Participants were workers aged 20 to 65 years at the time of the survey and were stratified by cluster sampling according to gender, age, and region of residence. After participants with insufficient responses were excluded, 27,036 individuals were enrolled. We then excluded 195 people infected with COVID-19 and 204 who were close contacts, meaning that 26,637 were finally available for analysis. This study was approved by the Ethics Committee of the University of Occupational and Environmental Health, Kitakyushu, Japan (Approval No. R2-079).

### Assessment of the 2020/2021 seasonal influenza vaccination

Regarding the 2020/2021 seasonal influenza vaccination, we asked participants “Did you receive this season’s influenza vaccine?,” to which they answered “Yes” or “No.”

### Assessment of sociodemographic factors

Regarding sociodemographic factors, we investigated gender, age (20–29, 30–39, 40–49, 50–59, and 60–65 years), marital status (single; divorced or widowed; married), education (junior high school or high school; vocational school or college; university or graduate school), annual household income (<2.00 million Japanese yen [JPY]; 2.00–3.99 million JPY; 4.00–7.99 million JPY; ≥8.00 million JPY), and underlying disease, for which we asked the question, “Do you have any disease that requires regular visits to the hospital or treatment?” Participants selected one of the following: “I do not have such a disease,” “I am receiving hospital visits and treatment as scheduled,” or “I am not receiving hospital visits and treatment as scheduled.” We rated the participants who answered, “I do not have such a disease” as “No” and the remaining two answers as “Yes.”

### Statistical analysis

Seasonal influenza vaccination rates were calculated for all variables. We set the presence or absence of 2020/2021 seasonal influenza vaccination as the dependent variable, and each aspect of sociodemographic factors consisting of gender, age, marital status, education, annual household income, and underlying disease as independent variables. Multivariate adjusted odds ratios (ORs) were estimated using multilevel logistic regression analysis. The multivariate model was adjusted for all covariates. Considering the influence of regional differences in the infection status of COVID-19, all analyses were performed by multilevel analysis nested by prefecture of residence. A *P* values of less than 0.05 were considered statistically significant. All analyses were performed using STATA Version 16 (StataCorp, College Station, TX, USA).

## Results

Table 1 shows the characteristics of the participants. A total of 26,637 people (13,600 men and 13,037 women) were analyzed. In each variable, 50–59 years old (8,909: 33.5%), married (14,794: 55.4%), annual household income of 4.00-7.99 million JPY (11,759: 48.2%), and university or graduate school (12,956: 48.6%) provided the largest numbers. There were 9,346 people (35.1%) with underlying disease. A total of 11,404 individuals (42.8%) received the 2020/2021 influenza vaccine.

**Table 1.** Characteristics of the participants

|  | Total<br>n=26637 |
| --- | --- |
| Gender |  |
| Men | 13600 (51.1%) |
| Women | 13037 (48.9%) |
| Age (years) |  |
| 20–29 | 1850 (7.0%) |
| 30–39 | 4762 (17.9%) |
| 40–49 | 7904 (29.7%) |
| 50–59 | 8909 (33.5%) |
| 60–65 | 3212 (12.1%) |
| Marital status |  |
| Single | 9034 (33.9%) |
| Divorced or widowed | 2809 (10.6%) |
| Married | 14794 (55.4%) |
| Education |  |
| Junior high or high school | 7240 (27.2%) |
| Vocational school or college | 6441 (24.2%) |
| University or graduate school | 12956 (48.6%) |
| Annual household income (JPY) |  |
| <2.00 million | 1685 (6.3%) |
| 2.00–3.99 million | 5487 (20.6%) |
| 4.00–7.99 million | 11759 (48.2%) |
| ≥8.00 million | 7706 (28.9%) |
| Underlying disease |  |
| No | 17291 (64.9%) |
| Yes | 9346 (35.1%) |
| Receiving 2020/2021 seasonal influenza vaccine |  |
| No | 15233 (57.2%) |
| Yes | 11404 (42.8%) |
| JPY, Japanese yen |  |

Table 2 shows the sociodemographic factors associated with receiving the 2020/2021 seasonal influenza vaccine. In the multivariate adjusted model, significantly more women than men were vaccinated (aOR 1.52, 95% CI 1.43–1.61). Compared with people aged 20–29, those aged 30–39 (aOR 0.88, 95% CI 0.79–0.99), 40–49 (aOR 0.68, 95% CI 0.61–0.76), 50–59 (aOR 0.64, 95% CI 0.57–0.71), and 60–65 (aOR 0.72, 95% CI 0.63–0.82) had lower vaccination rates. Married people were more likely to be vaccinated than single people (aOR 1.72, 95% CI 1.61–1.83), and more vaccinations were administered to individuals with higher levels of education, and those with higher annual household income. In addition, individuals with underlying diseases were also more likely to be vaccinated (aOR 1.55, 95% CI 1.47–1.64). (Table 2).

**Table 2.** Sociodemographic factors associated with receiving 2020/2021 seasonal influenza vaccine

|  | n | %<sup>a</sup> | Crude |  |  | Multivariate adjusted* |  |  |
| --- | --- | --- | --- | --- | --- | --- | --- | --- |
|  |  |  | OR | 95% CI | P value | OR | 95% CI | P value |
| Gender |  |  |  |  |  |  |  |  |
| Men | 5310 | 39.0 | 1.00 | – | – | 1.00 |  |  |
| Women | 6094 | 46.7 | 1.37 | 1.31–1.44 | <0.001 | 1.52 | 1.43–1.61 | <0.001 |
| Age (years) |  |  |  |  |  |  |  |  |
| 20–29 | 908 | 49.1 | 1.00 | – | – | 1.00 | – |  |
| 30–39 | 2294 | 48.2 | 0.97 | 0.87–1.08 | 0.619 | 0.88 | 0.79–0.99 | 0.031 |
| 40–49 | 3226 | 40.8 | 0.73 | 0.66–0.80 | <0.001 | 0.68 | 0.61–0.76 | <0.001 |
| 50–59 | 3558 | 39.9 | 0.71 | 0.64–0.78 | <0.001 | 0.64 | 0.57–0.71 | <0.001 |
| 60–65 | 1418 | 44.1 | 0.84 | 0.75–0.95 | 0.004 | 0.72 | 0.63–0.82 | <0.001 |
| Marital status |  |  |  |  |  |  |  |  |
| Single | 3233 | 35.8 | 1.00 | – | – | 1.00 | – |  |
| Divorced or widowed | 1113 | 39.6 | 1.16 | 1.07–1.27 | 0.001 | 1.33 | 1.21–1.46 | <0.001 |
| Married | 7058 | 47.7 | 1.62 | 1.53–1.70 | <0.001 | 1.72 | 1.61–1.83 | <0.001 |
| Education |  |  |  |  |  |  |  |  |
| Junior high or high school | 2609 | 36.0 | 1.00 | – | – | 1.00 | – |  |
| Vocational school or college | 2940 | 45.6 | 1.53 | 1.43–1.64 | <0.001 | 1.40 | 1.31–1.51 | <0.001 |
| University or graduate school | 5855 | 45.2 | 1.53 | 1.44–1.63 | <0.001 | 1.41 | 1.32–1.50 | <0.001 |
| Annual household income (JPY) |  |  |  |  |  |  |  |  |
| <2.00 million | 441 | 26.2 | 1.00 | – | – | 1.00 | – |  |
| 2.00–3.99 million | 2,034 | 37.1 | 1.65 | 1.46–1.87 | <0.001 | 1.60 | 1.41–1.81 | <0.001 |
| 4.00–7.99 million | 5,145 | 43.8 | 2.19 | 1.95–2.45 | <0.001 | 1.93 | 1.71–2.17 | <0.001 |
| ≥8.00 million | 3,784 | 49.1 | 2.76 | 2.45–3.11 | <0.001 | 2.28 | 2.01–2.58 | <0.001 |
| Underlying disease |  |  |  |  |  |  |  |  |
| No | 6861 | 39.7 | 1.00 | – | – | 1.00 | – |  |
| Yes | 4543 | 48.6 | 1.43 | 1.36–1.50 | <0.001 | 1.55 | 1.47–1.64 | <0.001 |

\*Adjusted for gender, age, marital status, educational background, annual household income, and underlying disease
<sup>a</sup>Row proportions for influenza vaccination
OR, odds ratio; CI, confidence interval; JPY, Japanese yen

## Discussion

The COVID-19 pandemic constitutes a unique situation in which the expectations for the efficacy of the 2020/2021 seasonal influenza vaccine included the prevention of not only influenza itself, but also the possibility of being treated as COVID-19. We investigated the effects of socioeconomic factors on seasonal influenza vaccine inoculation in this context. The results revealed that the influenza vaccination rate was higher among women, younger adults, married people, highly educated people, high-income earners, and those with underlying disease.

A review by Kini et al. reported that women were more likely to receive the seasonal influenza vaccine and that older people had higher vaccination rates than young people.^11^ In addition, it has reported that not being married was associated with non-vaccination, whereas higher levels of education and the presence of chronic diseases increased the likelihood of vaccination.^12,14^ Thus, other than age, the current results were consistent with previous reports regarding sociodemographic factors.

Previous studies have suggested various factors that may contribute to the higher vaccination rates observed among older people. Because older people are more likely to have chronic disease, vaccinations are often recommended by their physicians. Older people are also more likely to have had the influenza infection in the past, and to believe in the benefits of vaccination.^15,16^ In contrast, many young people have little knowledge about the risks of contracting influenza or about vaccine efficacy, and they often do not receive vaccine recommendations, have fewer opportunities to obtain such information, or ignore information that is provided.^15,17^ In addition, adverse reactions following vaccination, including local and systematic symptoms, have been more frequently reported in younger people, which has contributed to lower vaccination rates among young people.^11^

The results of the current study suggest that young people became more aware of the risk of contracting influenza during the COVID-19 pandemic, potentially resulting in higher vaccination rates. If contracting influenza, it was necessary to consult a medical institution or contact a health center in a situation whereby it was difficult to distinguish influenza from COVID-19, complicated by problems that occurred in the diagnostic process and results. At the time of the survey in Japan, where the number of infected people was smaller than in Western countries and the establishment of testing systems such as RT-PCR was delayed, it took time to make a definitive diagnosis and there was also prejudice against infected people.^12^ Thus, risk perception among young people may have increased. Risk perception has a great influence on vaccination intention. A meta-analysis of 34 papers, including studies on influenza vaccines, showed that the higher is risk perception, the greater is the number of vaccinations.^18^ Young people may have also become more aware of the effectiveness of the seasonal influenza vaccine. Vaccination decisions are typically made by comparing the positive and negative aspects of vaccination, such as the efficacy of the vaccine and the risks of side effects.^19^ An important aspect of the 2020/2021 seasonal influenza vaccine was that the perceived efficacy included preventing not only influenza itself but also the possibility of preventing COVID-19, as described above.^3^ Although young people are more likely to experience the side effects of the vaccine,^11^ it is possible that awareness of vaccine efficacy outweighed concerns about side-effects. We speculate that these changes in young people may have been influenced by information regarding vaccine efficacy being effectively provided to many young people through government announcements, media such as TV and the internet, and possibly in the workplace.

Regarding factors other than age, women, married people, highly educated people, high-income earners, and those with underlying diseases exhibited higher vaccination rates in the current study, which was similar to results reported in previous studies.^11,12,14^ These results suggest that the effects of these factors on seasonal influenza vaccination did not change, even in the unique context of the COVID-19 pandemic.

Overall, the annual influenza vaccination rate for the Japanese population is reported to be around 30%–40% for adults under the age of 65 years,^9,10^ and in this study conducted in December the vaccination rate was already slightly higher than was stated in these reports. This may be due to the relatively high risk of contracting seasonal influenza under the COVID-19 pandemic, resulting in increased perception of vaccine efficacy, particularly among young people. This result indicates that the seasonal influenza vaccination rate increased in young people who usually have a relatively low inoculation rate. This finding may have important implications. Education about the risk of infectious diseases is likely to have contributed to this result, and information about vaccine efficacy and the risk of side effects of the vaccine was effectively provided and understood. This finding may be useful for informing initiatives to improve vaccination rates in the future.

The main strength of the current study was the large sample size. However, two main limitations were involved in the current study. First, because we conducted an Internet survey, selection bias might have occurred. Although we stratified by cluster sampling according to gender, age, and region of residence at the beginning of the survey to reduce bias, the survey was intended for people registered as an online survey panel and was not representative of the general population. Second, regarding influenza vaccination, we asked subjects whether they were vaccinated between December 22 and 26 in 2020, when the survey was conducted; therefore, those who were vaccinated after that time and those scheduled to be vaccinated were not included. In particular, the 2020/2021 influenza vaccine has been in short supply in some areas and medical institutions owing to high demand for vaccination because of the influence of the COVID-19 pandemic, and in some cases, there was a long waiting time before inoculation. However, since vaccination started at the end of October in Japan, it was probable that those who had decided to be inoculated had collected information on supply shortages and made reservations and vaccinations early.

In conclusion, in a situation in which the rate of seasonal influenza vaccination increased during the COVID-19 pandemic, the relationships between seasonal influenza vaccination behavior and sociodemographic factors differed from the findings of previous studies in terms of age. Our results suggest that young people may have become more aware of the risks of contracting influenza during the COVID-19 pandemic, and of the effectiveness of the seasonal influenza vaccine. This increased awareness may have resulted in higher vaccination rates. We speculate that information interventions contributed to this result. We believe that these findings provide useful insight for improving the influenza vaccination rate.

## Data Availability

Data not available due to ethical restrictions

## Acknowledgments

We thank the current members of the CORoNaWork Project, in alphabetical order, are as follows: Dr. Yoshihisa Fujino (present chairperson of the study group), Dr. Akira Ogami, Dr. Arisa Harada, Dr. Ayako Hino, Dr. Chimed-Ochir Odgerel, Dr. Hajime Ando, Dr. Hisashi Eguchi, Dr. Kazunori Ikegami, Dr. Keiji Muramatsu, Dr. Koji Mori, Dr. Kosuke Mafune, Dr. Kyoko Kitagawa, Dr. Masako Nagata, Dr. Mayumi Tsuji, Dr. Rie Tanaka, Dr. Ryutaro Matsugaki, Dr. Seiishiro Tateishi, Dr. Shinya Matsuda, Dr. Tomohiro Ishimaru, Dr. Tomohisa Nagata, and Ms. Ning Liu. All members are affiliated with the University of Occupational and Environmental Health, Japan.

